# Factors associated with antenatal care service utilization among women with children under five years in Sunyani Municipality, Ghana

**DOI:** 10.1101/2021.02.27.21252585

**Authors:** Sekyere Stephen Owusu

## Abstract

**Background:** Maternal and neonatal mortality remains a public health burden around the globe most especially in developing countries. A well utilized antenatal care (ANC) is however among the identified interventions to reduce this burden of maternal and neonatal mortality rates. A lot of factors therefore predispose, enable and cause mothers to identify the need to utilize this service (ANC).

**Aims/objectives:** The aim of this study was to determine factors associated with the utilization of ANC service among postpartum mothers in the Sunyani municipality.

**Methods:** This study employed descriptive cross-sectional survey design. A semi-structured questionnaire consisting of demographic profile of the respondents, their knowledge about ANC services and the level of ANC utilization. Logistic regression analysis techniques and chi-square were used for the categorical variables to examine the associations between the dependent and independent variables. Data analysis was done using the Statistical Package for Social Science software (SPSS) version 22.

**Results:** Majority (87%) of postpartum mothers in the Sunyani municipality attended ANC at least once during their last pregnancy of which 95.6% had four or more visits and 77.1% initiated their ANC attendance within their first trimester. It was further observed that 97.3% of the mothers had good knowledge about ANC. Marital status and ANC knowledge were found to be significantly associated with ANC attendance. Husbands were found to be poorly involved in ANC services in the Sunyani municipality.

**Conclusion:** Marital status and ANC knowledge predisposes most mothers to utilize ANC services. Hence, health education activities about ANC services and its importance should be channeled more to areas where ANC utilization is low.

## CHAPTER ONE

### 1.0 BACKGROUND

#### 1.1 Introduction

The International human rights law includes fundamental commitments of states to enable women and adolescent girls to survive pregnancy and childbirth as part of their enjoyment of sexual and reproductive health rights and living a life of dignity (Human Rights Council, 2012). The World Health Organization (WHO) also envisions a world where “every pregnant woman and newborn receives quality care throughout their pregnancy, childbirth and postnatal periods” (Tunçalp et al., 2015, p.2). The basic materialization of this vision is only through the efforts of a well utilized antenatal care service, as the world health organization have demonstrated a positive relationship between ANC service utilization and pregnancy outcomes (World Health Organization, 2016).

Antenatal care (ANC) is a type of health service upkeep provided by skilled health professionals to pregnant women so as to ensure the best health condition of both the mother and the unborn baby throughout the pregnancy period (WHO, 2016). This service is composed of risk identification, prevention and management of pregnancy related or concurrent conditions, and health education and health promotion with essential interventions such as early identification and management of obstetric complications (pre-eclampsia and eclampsia), identification and management of sexually transmitted infections (HIV/AIDS, syphilis and others), tetanus toxoid immunization, and intermittent preventive treatment for malaria during pregnancy (IPTp) enshrined in it (WHO, 2016).

Regardless of the fact that about half of pregnant women globally received at least one prenatal care in the year 2013, most pregnant women persistently miss these indispensable services provided by antenatal care (Lincetto, Mothebesoane-anoh, Gomez, & Munjanja, 2013). Kuhnt and Vollmer (2017) found in their research that at least one ANC visit is associated with 1.04% points reduced probability of neonatal mortality and 1.07% points lower probability of infant mortality. Moreover, it was found in the same study that, at least one ANC visit is associated with 3.82% points reduced probability of giving birth to a low birth weight baby, 4.11% and 3.26% points reduced stunting and underweight probability respectively. A conclusion was therefore drawn from the same study that, currently, the existing and accessed ANC services in most low-income and middle-income countries are directly associated with improved birth outcomes and long term reduction of child mortality and malnourishment (Kuhnt & Vollmer 2017).

Similarly, according to Abbas et al. (2017), women who irregularly attends antenatal care during their pregnancy period stands at a higher probability of experiencing pregnancy complications such as preeclampsia, eclampsia and anemia. All these factors contribute to an increase in maternal mortality besides higher adverse birth outcomes of preterm birth, low birth weight and stillbirth. The world health organization specifies that, Maternal Mortality Ratio (MMR) which is simply the death of a woman whiles pregnant due to issues aggravated by the pregnancy is considered to be low if it is less than 100, moderate if it is between 100 and 299, high if it is between 300 and 499, very high if it is 500 and 999 and extremely high if it is equal to or higher than 1000 maternal deaths per 100 000 live births (WHO, 2015). In this regard, an analysis of about 183 countries and territories revealed an estimated 303 000 maternal deaths in the year 2015, yielding an overall MMR of 216 (UI 207 to 249) maternal deaths per 100 000 live births of which low resource settings alone contributed 99% to this record (WHO, UNICEF, UNFPA, Group, & UNPD, 2015).

For WHO regions, the overall MMR in developing regions is estimated at 239 (UI 229 to 275), which is roughly 20 times higher than that of developed regions, where it is just 12 (UI 11 to 14) (UNICEF, 2015). Sub-Saharan Africa has a very high MMR with a point estimate of 546 (UI 511 to 652) (UNICEF, 2015). According to the World Health Statistics (2017), there were 195000 maternal deaths in Africa in the year 2015. Ghana in the same year (2015) also made a record of 2800 maternal deaths representing MMR of 319 (UI 216 to 458) (United Nations Population Division, 2015).

Between the periods of 1990 and 2015, the global maternal mortality ratio (MMR) decreased by 44%, from 385 to 216 maternal deaths per 100,000 live births (Maternal Health Task Force, 2017). Despite this progress, the world still fell far short of one of the then Millennium Development Goals target of a 75% reduction in the global MMR by 2015 (MDG 5 target 5.a). Notwithstanding, maternal mortality reduction remains a priority under Goal 3: “Ensure healthy lives and promote well-being for all at all ages” in the new Sustainable Development Goals (SDGs) agenda through 2030 (Tangcharoensathien, Mills & Palu, 2015). Specifically, target 3.1 of SDG 3 aim to reduce the global maternal mortality ratio to less than 70 per 100,000 live births by 2030 and target 3.2 aims to end preventable death of infants and children under 5 years of age, with all countries aiming to reduce neonatal mortality to at least as low as 12 per 1000live births and under-5 mortalities to at least as low as 25 per 1000live births by 2030 (Costanza et al., 2016). Target 3.2 of the aforementioned goal (SDG) can be achieved when antenatal care is being utilized together with child welfare services.

This paper was therefore intended to examine some of the various factors associated with ANC service utilization specifically in the Sunyani municipality among postpartum mothers to help ignite the maximum utilization of the service (ANC) for healthy pregnancy experience and outcomes in this municipality which will indirectly cause a reduction in the global, national and regional maternal mortality ratio (MMR) and contribute to the attainment of the third Sustainable Development Goal (SDG 3: *“Ensure healthy lives and promote well-being for all at all ages”*) as well make the vision of WHO (*“a world where every pregnant woman and newborn receives quality care throughout their pregnancy, childbirth and postnatal periods”*) becomes a reality.

#### 1.2. Problem statement

Pregnancy related deaths and disorders remains inadmissibly high at the global front claiming millions of women and infants’ life (WHO, 2016). It is a recommendation by the world health organization that, all pregnant women should have at least four antenatal visits throughout their pregnancy period (WHO, 2016). However, in Sumankuuro et al.’s (2016) study, it was reported that government of Ghana’s effort to increase uptake of ANC significantly continue to be awkward in rural Ghana. Globally, while 86 percent of pregnant women access antenatal care with skilled health personnel at least once, only three in five (62 percent) receives at least four antenatal visits which is even worse in countries with high maternal mortality rates such as sub-Saharan Africa (51.9%) and least developed countries including Ghana (45.8%) (UNICEF, 2017). Government of Ghana in an attempt to reduce maternal mortality introduced free maternal health service policy to break financial barriers of access to maternal care services. Despite this effort by the government of Ghana, facility based deliveries continue to be low, attributed to the poor quality of antenatal care which prevents pregnant women from giving birth at health facilities (Atinga & Baku, 2013).

A critical look at the ANC coverage in Ghana between the periods of 2011 to 2016 revealed that, Ghana during the aforementioned period experienced a sharp decline in ANC coverage in the order of 98.2% - 92.1% - 90.8% - 87.3% - 84.5% and 84.1% respectively (Ministry of Health, 2016). Also, according to data obtained from the Sunyani district health directorate, the case of Sunyani municipality is of no difference in terms of the continuous declination of ANC coverage as the district between 2014 and 2017 experienced a sharp decline of ANC coverage from 133.2% - 123.8% - 123.1% and 115.6% respectively. As a result, the district between the same period (2014 – 2017) suffered an increasing order of maternal mortality rate per 100,000 live births ranging from 262/100,000 – 271.6/100,000 – 283/100,000 and 296/100,000 respectively. The World Health Organization stated that “half a million mothers and 10.6 million children will die globally in each year if governments do not increase their efforts to reduce maternal and child deaths” (Ian, 2005 p.3). It is therefore important to investigate into the factors associated with ANC utilization to help with the implementation of necessary policies which can help deal with maternal deaths and help produce healthy babies.

#### 1.3. Justification

According to the United Nations Inter-Agency Group for Child Mortality Estimation, maternal and child mortality rates are among the most important indicators of child health, nutrition, implementation of key survival interventions, and the overall social and economic development of a population (United Nations Inter-Agency Group, 2016). The health of pregnant women and their unborn babies cannot be underestimated, and as stated by the UN Secretary-General Ban Ki-moon cited in an overview document on WHO guideline on antenatal care that “To achieve every Woman every Child vision and the Global Strategy for Women’s, Children’s and Adolescents’ Health, we need innovative, evidence-based approaches to antenatal care” (WHO, 2016 p.45). Similarly, Dr. Janet Campbell, a passed civil servant in UK stated as early as 1929 that “the first requirement of maternity service is effective supervision of the health of the woman during pregnancy”. This conversely is the kind of service provided by antenatal care.

Therefore as public health agencies together with the world health organization, Ghana health service and other non-governmental agencies continually puts up efforts and strategies in an attempt to make life in communities, regions, countries and the world at large become enjoyable, maternal health with a focus on ANC cannot be exempted because the death of a pregnant woman poses much burden and loss to the affected family and the community since two lives can be lost in the process (both the woman and the foetus). Issues of maternal health focusing on antenatal care therefore requires prompt attention in the field of health and research. While a lot of studies have been conducted in neighboring countries and other regions in Ghana concerning antenatal care service utilization, no known specific studies have been done to identify the factors associated with ANC service utilization in Sunyani municipality of Ghana and where postpartum mothers were sampled. This knowledge gap could potentially hamper effective planning and delivery of reproductive and maternal healthcare services. Acquiring knowledge about the factors associated with ANC service utilization could therefore constitute an important step of considering measures that will help increase the utilization of ANC services generally and specifically in the Sunyani municipality of Ghana. This study therefore seeks to fill this lacuna and add on to existing literature by investigating the factors associated with ANC service utilization among postpartum mothers in the Sunyani municipal district.

#### 1.4. Aim of the Study

This study was aimed at determining factors associated with antenatal care (ANC) service utilization in Sunyani municipality of Ghana.

#### 1.5. Specific Objectives

1. To determine antenatal care service utilization among postpartum mothers in Sunyani municipality.
2. To determine the effect of socio-demographic factors (Age, marital status, economic status, level of education, place of residence etc.) on antenatal attendance.
3. To assess the knowledge level of postpartum mothers about antenatal care in the Sunyani municipality.
4. To determine the relationship between knowledge and antenatal care service utilization.

## CHAPTER TWO

### 2.0 LITERATURE REVIEW

#### 2.1 Introduction

Due to the complicated nature of pregnancy, it is associated with a lot of problems and disorders such as pregnancy induced hypertension (eclampsia), obstructed labor and others which poses threat to the developing foetus and the pregnant mother. This lead to the inception of the antenatal care model under the safe motherhood program in the early 1900s which has since been under intense scrutiny to ensure its optimal quality and utilization (WHO, 2016). For example, the earlier model known as Focused Antenatal Care approach (FANC) was reviewed and it was revealed that the limited number of visits (four visits) was associated with an increased perinatal death, maternal mortalities and low satisfaction rate of pregnant mothers (Vogel et al., 2013; Dowswell et al., 2010). The number of visits was then increased from four (4) to eight (8) and making it more interactive than a mere visit hence the term *‘visits’* was changed to *‘contacts’* which lead to the recent WHO antenatal care model since November, 2016 (Tunçalp et al., 2017).

#### 2.2 Antenatal care service utilization

After the failure of the Focused ANC to address maternal mortality and perinatal deaths (Vogel et al., 2013; Dowswell et al., 2010), ANC utilization since then is measured against WHO’s new recommendation of having at least four contacts throughout a woman’s pregnancy period and initiating it within the first trimester (WHO, 2016). There has been good progress in ANC utilization in developing countries where the majority (80%) of women receive at least one ANC check-up (Wang, Alva, Wang, & Fort, 2011). Similarly, in a range of LMICs from 2011 to 2017, nearly all women had at least one ANC visit and a significant increase in the proportion of women who received at least four visit across all sites (Tikmani et al., 2019).

In sub-Saharan Africa, nearly three quarters (72%) of women initiate their first ANC check-up after the first trimester of pregnancy (WHO, 2016). Also according to Yamba (2018), about 98% of Sierra Leone women between 2009 and 2013 attended ANC during their last pregnancy where 89% had four (4) or more visits. To add on, a study by Abubakari and Abiiro (2018) among nursing mothers attending postnatal and child welfare clinics in three districts in Northern Ghana, it was found that approximately 81% of their study respondents have visited ANC four time or more with a coverage of over 99%. Also, Adu et al. (2018) analyzed data from the 2014 Ghana Demographic and Health Survey and found that irrespective of respondents’ marital status, majority of them had four or more antenatal visits (89.2%), while very few had less than four visits (10.8%).

In other literatures however, ANC underutilization is highly observed. In Nigeria for instance, the prevalence of ANC underutilization according one study is found to be 46.5% (Adewuyi et al., 2018). Per the WHO, only a small proportion (40%) of pregnant women in LMICs countries have attended the minimum four ANC visits (WHO, 2016). Moreover, despite nearly universal coverage of ANC in Rwanda, only 25% of pregnant women start ANC within the timeframe recommended by WHO (Mkandawire et al., 2019). As well, findings from Amoah et al. (2016) shows that, out of the 85% of their respondents who had at least one ANC visit in Ghana during the time of their study, only 31.7% had their first visit during their first trimester.

#### 2.3 Factors associated with Antenatal Care Service utilization

According to Anderson and Newman’s model of health service utilization, access to and use of a particular health service is a function of three main characteristics which includes predisposing factors, enabling factors and need factors (Anderson & Newman, 1973)

Predisposing factors: These take into consideration the socio-demographic and socio-cultural characteristics of the individual that exist prior to their medical condition (pregnancy) which will permit an individual to utilize a health service (ANC) more than others (Andersen & Newman, 1973). According to Andersen, Davidson and Baumeister (2007) individual predisposing factors include the demographic characteristics of age and sex as biological imperatives, social factors such as education, occupation, ethnicity and social relationships (e.g., family status), and mental factors in terms of health beliefs (e.g., attitudes, values, and knowledge related to health and health services)

Enabling factors: The enabling factors according to Anderson and Newman (1973) is considered to be the logistical aspects of health care utilization which takes into consideration the occupational status of the individual, income level, availability of health service, long waiting time, distance etc.

Need factors: The most immediate cause of health service use, from functional and health problems that generate the need for health care services. This also takes into account how people view their own general health and functional state, as well as how they experience symptoms of illness, pain, and worries about their health and whether or not they judge their problems to be of sufficient importance and magnitude to seek professional help (Andersen, Davidson, & Baumeister, 2007).

In a systematic review to identify the frequency of Andersen’s Behavioral Model (BM) use, it was found to be extensively used in studies investigating the use of health services but with a substantial difference in the variables used in the various categories. The majority of the reviewed studies included age (N=15), marital status (N=13), gender/sex (N=12), education (N=11), and ethnicity (N=10) as predisposing factors and income/financial situation (N=10), health insurance (N=9), and having a usual source of care/family doctor (N=9) as enabling factors for need factors, most of the studies included evaluated health status (N=13) and self-reported/perceived health (N=9) as well as a very wide variety of diseases (Babitsch, Gohl, & von Lengerke, 2012).

##### 2.3.1 Knowledge and perception as predisposing factors for ANC utilization

Knowledge about antenatal care service and the awareness of its tremendous benefits to pregnant women and their unborn baby is believed to be a major influential factor which triggers the decision of pregnant mothers to use ANC services. Many studies have proven it that, most pregnant women who do not use ANC is due to the lack of awareness and knowledge about the important provisions of ANC services.

For example, in a qualitative study to identify why women in low and middle income countries do not use ANC services, some pregnant mothers stated that, pregnancy is a natural process and hence does not require any further attention or care (Finlayson, & Downe, 2013). In addition, the perception of some individuals including some pregnant women regarding hospital attendance and health service utilization is that, health facilities are only made for those with disease conditions or disorders with apparent signs and symptoms. Birmeta, Dibaba and Woldeyohannes (2013)’s study confirms the above avowal where they found in their study that, a significant percentage of pregnant women in central Ethiopia did not attend ANC because they see pregnancy not to be a disorder hence there is no need to seek for health care when they are pregnant. In this regard, ignorance becomes the major factor which steals the minds of such women to access and utilize health service including ANC services. Baruah and Beeva (2016)’s work also adds on to the above discernment where they also found that some women did not attend antenatal care because they felt no need of seeing healthcare in times of pregnancy.

Besides, a study conducted in Tanzania which was aimed at establishing the association between knowledge of danger signs during pregnancy and subsequent health care seeking behavior, it was found that those who recognized danger signs were more willing to visit health facilities than those who were not knowledgeable about any danger signs of pregnancy (Mwilike et l., 2018). This shows that, mothers who attends ANC when pregnant also stands a higher chance of utilizing other health services due to the education they will receive at ANC sessions. A cross sectional study conducted in Western Jamaica among pregnant women depicts that women’s knowledge of ANC services is associated with the number of ANC attendance with those having good knowledge having four or more visits compared to those with poor knowledge (Respress et al., 2017). In addition, Ogboghodo, Adam, Omuemu and Okojie (2019) found good knowledge on maternal health issues to be a major predictor for skill birth attendance (SBA). In a study done in Manipur, it was observed that majority (97.9%) of postpartum mothers knew that pregnant women need to go for antenatal check-up but only 55.2% knew correctly about the minimum antenatal check-up during pregnancy (Laishram, ThounaojamUd, Mukhia, & Sanayaima, 2013). However, Kaur and colleague (2018) observed that 52% of mothers with infants in an urban area of Amritsar, Punjab knew the importance of ANC visits of which 89% of the mothers were able to tell the correct number of minimum ANC visits a pregnant woman should have throughout her pregnancy period. In the Ghanaian perspective regarding knowledge and perceptions about ANC services, one study conducted in the Ashanti region of Ghana revealed that, about 14% of pregnant women were having no knowledge about what antenatal care entails whiles 17% and 69% however had poor and fair to very good knowledge on ANC services respectively (Oppong, 2008).Yet, a study by Ogunba and Abiodun (2017) among mothers with infants (postpartum mothers) in South western Nigeria found no significant relationship between knowledge and the attendance of antenatal clinic.

That said, one study suggests that, there is no significant association between educational level of pregnant mothers and their level of knowledge about antenatal services (Maluleke, 2017).

##### 2.3.2 Financial difficulties as enabling factors for ANC utilization

Due to the underutilization of ANC services especially in most developing countries per WHO recommendations of minimum visits, many studies have been put forward to answer the why question for this low patronage. Among some of the factors found to be associated with antenatal care service utilization is financial difficulties. This can be in terms transportation cost or services charges (Titaley, Hunter, Heywood, & Dibley, 2010). Due to the poverty level in most developing countries as compared to the developed countries, financial inability hinders the rate at which health services is been accessed and utilized including antenatal care services leading to the double burden of disease and a mind blowing mortalities including maternal mortalities. In Nigeria, the two leading factors known to be associated with ANC service utilization according to one study from the perspective of the non-users includes getting money to go and Farness of ANC service (Fagbamigbe & Idemudia, 2015). These two factors can be merged and classified as financial powerlessness because if the individual is financially capable, distance will not be a barrier if only there is an availability of transport system. This result is however in agreement with that which was found in Ghana as cost, and distance influenced about 49% of pregnant women not to use ANC services (Asundep et al., 2013).

##### 2.3.3 Demographic features as predisposing factors for ANC utilization

Among the sociodemographic factors which influence antenatal care utilization includes; maternal age, educational level, parity, place of residence etc. Due to the Stigma associated with teenage pregnancy in some countries and societies, teenagers who becomes pregnant at their age feels reluctant and shy to disclose their pregnancy early. Maternal age has however been marked as a factor associated with antenatal care service utilization. For instance, according to Doku, Neupane and Doku (2012), Women aged 25-34 years old have higher probability of having early ANC visit compared to those aged 15-24 years old and also the older a woman, the more likely it is that she will have a delivery assisted by trained health personnel. This finding is however consistent with what was found by Chubike and Constance (2013) where mothers between the ages of 25 and 34 had the highest ANC attendance, institutional delivery and skilled attendant. Moreover, Joshi et al. (2014) also found older age to be positively associated with ANC attendance. Recently, Adewuyi et al. (2018) also observed that maternal age is associated with ANC utilization among Nigerian women and also a significant difference between rural residents and urban residents as far as ANC utilization is concerned. The prevalence of ANC underutilization among those in rural areas was high (61.1%) compared to 22.4% among those in the urban settings of which lack of companionship to health facility and getting money to go was found to be some of the reasons for this disparity.

Considering marital status as a predictor for ANC utilization, a study conducted in rural Ghana found that, cohabiting women and unmarried women were 43% and 61% respectively less likely to have attended ANC at least four times relative to married women (Sakeah et al., 2017). In addition, findings from Ziblim, Yidana and Mohammed (2018) shows a significant association between marital status and ANC utilization. Furthermore, it has been found that women who are single, divorced, widowed or separated are at a higher risk of poor utilization of ANC services as compared to married women according to a study conducted among recently delivered women in Rwanda (Rurangirwa et al., 2017). But according to Nuamah et al. (2019), there is a significant association between marital status and ANC utilization with cohabiting mothers having a higher odds of attending ANC compared to those who are married in the forest belt of Ghana. Also findings from Kparu (2016)’s study among adolescents in Ghana suggest that, comparing marital status and number of ANC visit, 66 (58.9%) of adolescent who were single made ANC visit of 4 times or more, cohabiting 16 (43.2%), married 15 (48.4%) indicating no significant association between marital status and ANC attendance.

Educational level is also another socio-demographic factor associated with the rate at which pregnant women utilize health services including antenatal care services. Although some studies suggest that there is no significant association between mother’s educational level and their knowledge about ANC services (Maluleke, 2017; Adu, 2018) other studies still suggest that literacy accounts for mother’s willingness to attend and use antenatal services. Considering a study by Baruah and Beeva (2016), about 50% literate mothers had between one and three (1 and 3) ANC contacts compared to about 40% illiterate mothers who had the same number of ANC contacts (between one and three). In addition, the three indicators for maternal health service utilization (Antenatal services, Skilled birth attendance and postpartum service) increased sharply with increased maternal education where further analysis indicated that women with less education in the lowest wealth quintile continue to use traditional medicine for maternal and child health services in Eritrea (Habtom, 2017). In some societies, due to husband’s autonomy in decision making, their level of education have been found to be associated with their wife’s health care seeking behavior including their maternal health. Baruah and Beeva (2016) observed that, women whose husband had a post-secondary education showed a positive attitude towards their maternal health including ANC utilization.

All in all, despite the consistent pattern of the observed factors associated with ANC utilization (knowledge, cost, distance, and demographic characteristics) from many of the reviewed literatures, conclusion cannot be drawn that these are the only associated factors. Manda-Taylor, Sealy and Roberts (2017) found other multiple factors such as attitudes toward pregnancy, hospital inefficiencies, lack of spousal support and conflicting ANC promotion messages to be also associated with ANC utilization.

#### 2.4 Including Husbands in Antenatal Care Services

The inclusion of men in maternal and safe motherhood services is increasingly recognized as an important determinant of women’s access to needed care in many low-income settings, including Ghana. In societies where patriarchal norms are dominant, men are often major decision-makers for the family, hence decisions around when, where and even if a woman should have access to healthcare often are made by their husbands. Women who receive education together with their husbands at ANC are more likely to attend postpartum services and also prepare more effectively for their pregnancies as compared to women whom received the education alone (Ganle, & Dery, 2015; Mullany, Becker, & Hindin, 2006).

Still and all, it has been recently found that, interventions to engage men in ANC services is associated with improved antenatal care attendance, skilled birth attendance, facility birth, postpartum care, birth and complications preparedness and improved maternal nutrition of pregnant women and also improves male partner’s support for wife as well increases couple communication and joint decision making, with ambiguous effects on women’s autonomy (Tokhi et al., 2018). In peri-urban Gulu district, in Northern Uganda the proportion of male partners that participated in at least one ANC visit was 65.4%. Although most men (60.5%) attended not more than two ANC visits, their willingness to attend subsequent ANC visits with their spouses was observed to be at a higher percentage of 93.7% (Tweheyo, Konde-Lule, Tumwesigye, & Sekandi, 2010).

However, evidence suggest that although most men are aware of the importance and the benefits of their involvement in maternal health care issues, their involvement level still falls below expectation (Ganle & Dery, 2015). The four major barriers to men’s involvement in maternal health services as identified by one study include: gender role and gender norm issues (perceptions that pregnancy care is a female role while men are family providers), negative socio-cultural beliefs such as the belief that men who accompany their wives to receive ANC services are being dominated by their wives, health services factors such as unfavorable opening hours of services, poor attitudes of healthcare providers such as maltreatment of women and their spouses and lack of space to accommodate male partners in health facilities as well as high cost associated with accompanying women to seek maternity care (Ganle & Dery, 2015).

According to Kwambai et al. (2013), the few percentage of men whom involve themselves in issues of maternal health are motivated by such factors as: to have a first-hand information about their wife and the unborn baby as some men believe that their wife may either distort or withheld some information after clinic visit and also to remind their spouse to follow the doctors’ instructions. Health providers denying services to women attending antenatal care without their partners, fast-tracking service to men attending antenatal care with their partners, and providing education and community sensitization are some of the available strategies adopted by some countries to help augment men involvement in maternal health services (Peneza & Maluka, 2018).

## CHAPTER THREE

### 3.0. MATERIALS AND METHOD

#### 3.1 Study Site Description

Sunyani municipality is one of the twenty-seven districts in the Brong Ahafo region. The municipality was established on 10th March, 1989 by a legislative instrument (LI) 1473.This was the period Ghana adopted the District Assembly concept. The overall goal is to accelerate growth and development in the Municipality.

The Sunyani Municipal district covers a total land area of 506.7 Km². It is located at the center of Brong Ahafo Region lying between Latitudes 70 20’N and 70 05’N and Longitudes 20 30’W and 2010’W. It is bordered on the north by Sunyani West District; west by Dormaa East District south by Asutifi District to the South and east by Tano North District. The population of the Sunyani municipality is about 123,224, made up of 61,610 males and 61,614 females. The municipality’s population represents 5.3% of the region’s total population. The population growth rate of the Municipality is estimated at 2.3%.

In relation to healthcare delivery, the municipality has a regional hospital, which caters for the inhabitants of the municipality and also serves as a referral point for the region. There is also a municipal hospital. There are three (3) health centers, one (1) mission hospital, two (2) private hospitals, three (3) Quasi clinics; Police Clinic, Prisons Clinic and thirteen (13) private Clinics, five (5) school clinics and three (3) maternity homes in the municipality. In all, there are thirty-four (34) health facilities in the municipality and out of these, eighteen (18) facilities provide antenatal and post-natal services for the community. Family planning services are provided in 21 out of the total of 34 health facilities in the municipality (Ghana statistical services, 2014).

#### 3.2 Study Population

This study was done among postpartum mothers who are within their reproductive age (15 – 49years), have delivered at least once in the last five years and is present at the Child Welfare Clinic for growth monitoring and promotion service for their babies.

#### 3.3 Inclusion and Exclusion Criteria

In order to avoid bias and obtain a credible data, the following criteria was set to give a boundary as to who qualifies to partake in the study. Mothers who falls within the ages of 15 – 49years and have delivered at least once in the last five years were included in the study. These group is believed to have experienced pregnancy before and have passed through the antenatal care stage of maternal health services and hence are the right people to be asked questions pertaining to pregnancy and antenatal care. Parental consent and child assent was sort for respondents below the age of 18years whom their mother/guardian accompanied them to the CWC unit. Respondents who were below the age of 18years and came to the CWC unit alone were not included due to ethical issues. Mothers who are above 18years but refused to give their consent were excluded from the study as well as those below the age of 18years whose parent/guardian refused to consent for their participation in the study. In addition, any woman within the defined age group, have not given birth before but was present at the CWC unit on behave of a friend or relative was excluded from this study.

#### 3.4 Study Design

This study employed the cross sectional survey design, using quantitative data collection and analysis methods. Cross-sectional studies provide a ‘snapshot’ of the outcome and the characteristics associated with it, at a specific point in time (Levin, 2006). Because this study aimed at identifying factors associated with ANC service utilization, a cross-sectional study design was appropriate as the design enables data to be collected on individual characteristics at the time of the study alongside information about the outcome, and association between individual characteristics and the outcome of interest.

#### 3.5 Sample Size Determination

The sample size of a study is a section of the population that is drawn to make inference or projections to the general population. The sample size for this study was calculated using the Cochran’s (1977) formula:

Sample size, 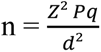 where;

**Z** being the confidence limits which in this study was 95% level of confidence and 1.96 as critical value.

**P** as the assumed prevalence / proportion of the dependent variable; According to a report by the Ghana demographic and health survey, large proportion of pregnant women in Ghana (87 percent) had four or more antenatal care visits for their most recent live births (GSS, 2014).

**Q** as the acceptable deviation from the assumed proportion (1-0.87 = 0.13).

**D** as the margin of error around p estimated as 0.05 in this study.

Therefore, 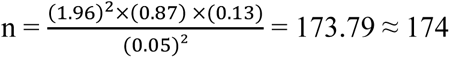

Adding 5% of 174 mothers (thus 174 × 0.05 = 8.7 ≈ 9), to compensate for errors during data collection, a total sample size of 183 (thus 174 + 9 = 183) mothers were recruited for the study.

#### 3.6 Sampling Method

This study employed the convenience sampling method. This is because the target population for the study (Post-partum mothers) are not always available at the child welfare clinic but their availability is influenced / determined by the EPI schedule. However, the Sunyani municipal health directorate was consulted to identify the various community health nurses who were on rotation to the various sub-districts and communities for outreach services on child welfare and know their schedules for the various communities during the period allocated for data collection. The data collectors were then assigned to the various community health nurses to the CWC units of the various communities for data collection. At each CWC unit, mothers who after explaining the purpose, benefits and risk of the study gives their consent to participate in the study were interviewed after they have received child welfare service for their child.

#### 3.7 Data Collection Procedure

A well-structured questionnaire comprising of both close ended and open ended questions were used to collect data from mothers at the Child Welfare Clinic of the various health facilities and outreach points where child welfare services are rendered. The questionnaires were designed in English language, but the questions were asked and explained in both English and the local dialects (i.e. *Bonu* and *Twi*). This ensured better understanding for mothers who have challenges with speaking the English language. The questionnaires were sectioned into A, B, and C where section A captured demographic data of the respondents and sections B and C captured data about knowledge and ANC utilization respectively. Knowledge level about ANC was measured using a 5-point Likert scale ranging from 1(one) to 5 (five). Four trained data collectors together with the researcher did the data collection. The overall aim of the study was explained vividly to the data collectors during the training session. The questionnaires were pretested at the Sunyani municipal hospital CWC unit with respondents of the same characteristics but were not included in the study. Data collection was done by distributing the questionnaires to mothers who were able read and understand the questions to write to fill the questionnaire on their own whiles the data collectors were still around to explain any ambiguities. For mothers who were unable to read and write, the questionnaire was filled by the data collectors whiles they ask the respondents the questions on the questionnaire.

#### 3.8 Data Analysis

Data from the field were edited, and checked for completeness by the researcher before data entry was done. Data was entered into EpiData manager and was exported into the Statistical Package for Social Sciences (SPSS) software version 22 for analysis. Results were displayed in tables and graphs according to the study variables. Bivariate analysis was done to test for association using Pearson’s correlation coefficient, Fisher exact test (for 2×2 tables where some of the expected counts are less than 10) and Likelihood Ratio (for bigger tables where 20% or more of the expected count is less than 5) in order to find out the relationship between the independent and dependent variables. P-value of less than 0.05 (i.e. p<0.05) was set as the significance level for the analysis.

Logistic regression model was then applied to variables which were significant at the bivariate analysis level to find out the strength of the association. Both simple and multiple logistic regression analysis were performed in order to get the crude and adjusted odd ratios respectively. The strength of the association for each independent variable was based on the odd ratios and the 95% confidence interval, while holding other factors constant. However, only the variables found to be significant for multiple logistics (adjusted odds ratios) were discussed in the study.

A reliability analysis was carried out on the ANC knowledge items to check for internal consistency using Cronbach’s alpha which gave an, α = 0.70. Most items appeared to be worthy of retention, resulting in a decrease in the alpha if deleted. Responses for the Likert scale were however recoded into ‘YES’ or ‘NO’. All responses of strongly agree and agree were recoded as *‘yes’* and all responses of no opinion to strongly disagree were recoded as *‘no’*. Median was computed as a measure of central tendency (what most respondents believe/think) to determine the knowledge level of respondents about ANC services. A descriptive frequency analysis of the newly computed/generated variable (ANC Knowledge level) showed and grouped respondents’ knowledge into either good or poor taking into consideration the interquartile rage (IQR) as a measure of spread/dispersion of responses.

Utilization of ANC was measured using the using parameters;

a. ANC attendance
b. Number of contacts (visits) per WHO recommendation
c. Gestational age at first contact (visit) per WHO recommendation.

#### 3.9 Ethical issues

Ethical clearance was sought from UHAS Research Ethics Committee (UHAS-REC) with a protocol identification number UHAS-REC A.4 [150] 18-19. Permission to proceed with the study was obtained from School of Public Health, University of Health and Allied Science, the District Health Management Team (DHMT), Hospital Management Team (HMT) as well as the Sunyani Municipal Health Directorate. An informed consent from respondents with clear explanation of procedure were also sought. Before issuing the questionnaires, all the respondents were presented with the option of declining to answer any of the questions. Furthermore, respondents were made aware of the fact that they may withdraw from the study at any point in time. Respondents’ safety was ensured and confidentiality of responses were also ensured.

#### 3.10 Limitations

The convenient sampling method which was employed in this study is prone to sampling bias. This however made the study likely to encounter systematic bias (difference in the results obtained from the study compared to the results obtained when the entire population is used) limiting the generalizability of the findings to the entire population. Also because respondents have to recall their experiences during their last pregnancy, recall bias was a limitation for this study. To reduce the occurrence of such limitations, 5% of the total sample size was computed and added to the calculated sample size. Also questions which respondents have to recall were all simple questions like *“did you attend ANC during your last pregnancy?”* and was framed in a way which will facilitate easy recall.

## CHAPTER FOUR

### 4.0 RESEARCH FINDINGS AND DISCUSSION

#### 4.1 Research Findings

A total sample size of 183 was calculated for the study using Cochran’s statistical formula for sample size calculation. However, a total of 182 data on respondents were analyzed for the study.

##### 4.1.1 Socio-demographic characteristics of respondents

Results in table 1, it is shown that majority 101 (56.4%) of the respondents are between the ages of twenty (20) and thirty (30) years with a mean (SD) age of 28.56 (SD = 5.99years). Marital status of the respondents indicates that most 151 (83.0%) of the respondents are married. In terms of employment status, most of the respondents 123 (69.1%) are self-employed. However, those who are unemployed 30 (16.9%) outweighed those who are government employed 25 (14.0%). Respondents place of residence seems to be almost balanced between urban 76 (41.8%) and rural 70 (38.5%) with the remaining 36 (19.8%) residing in peri-urban settings.

**Table 1.0.**
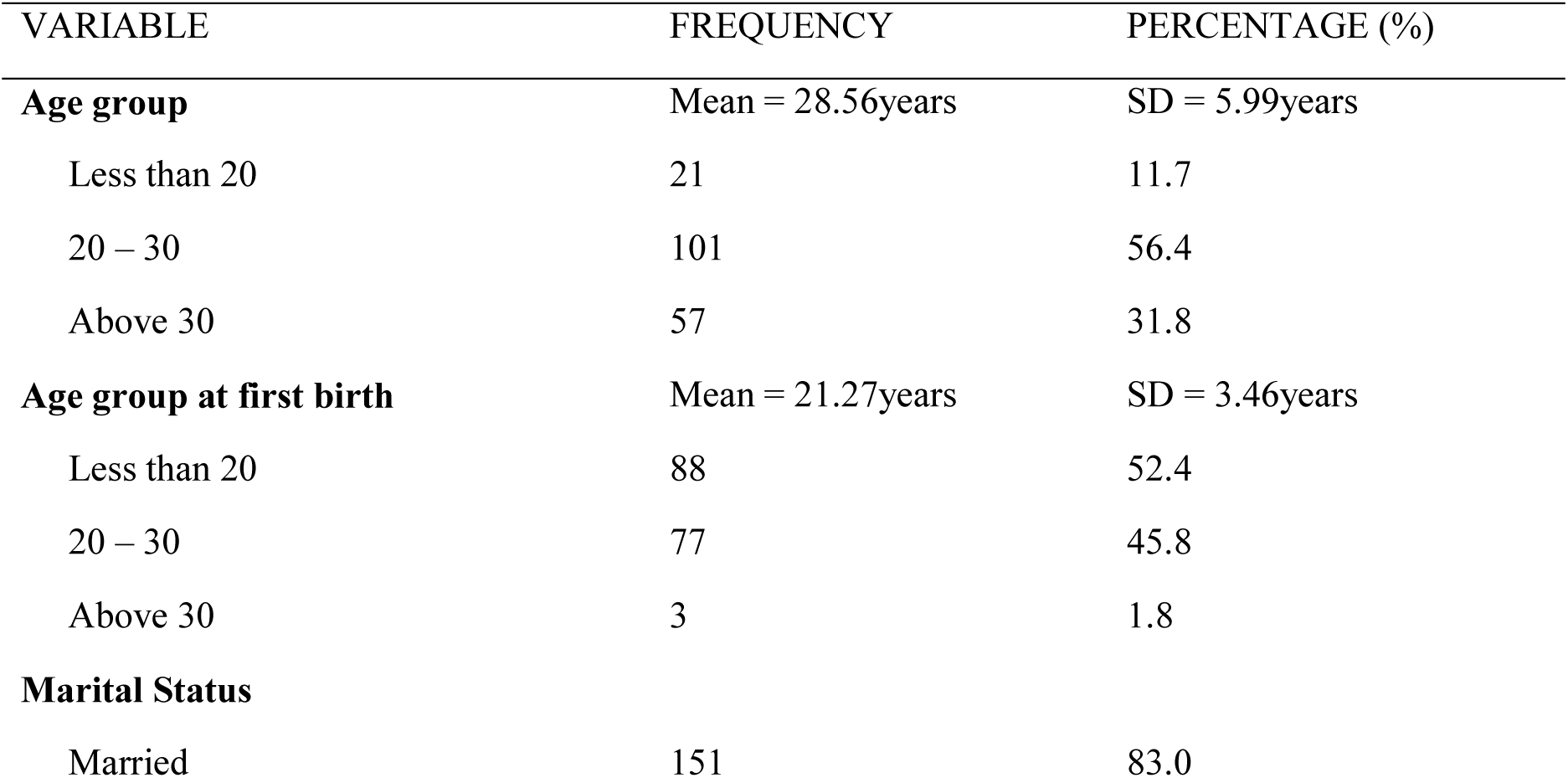
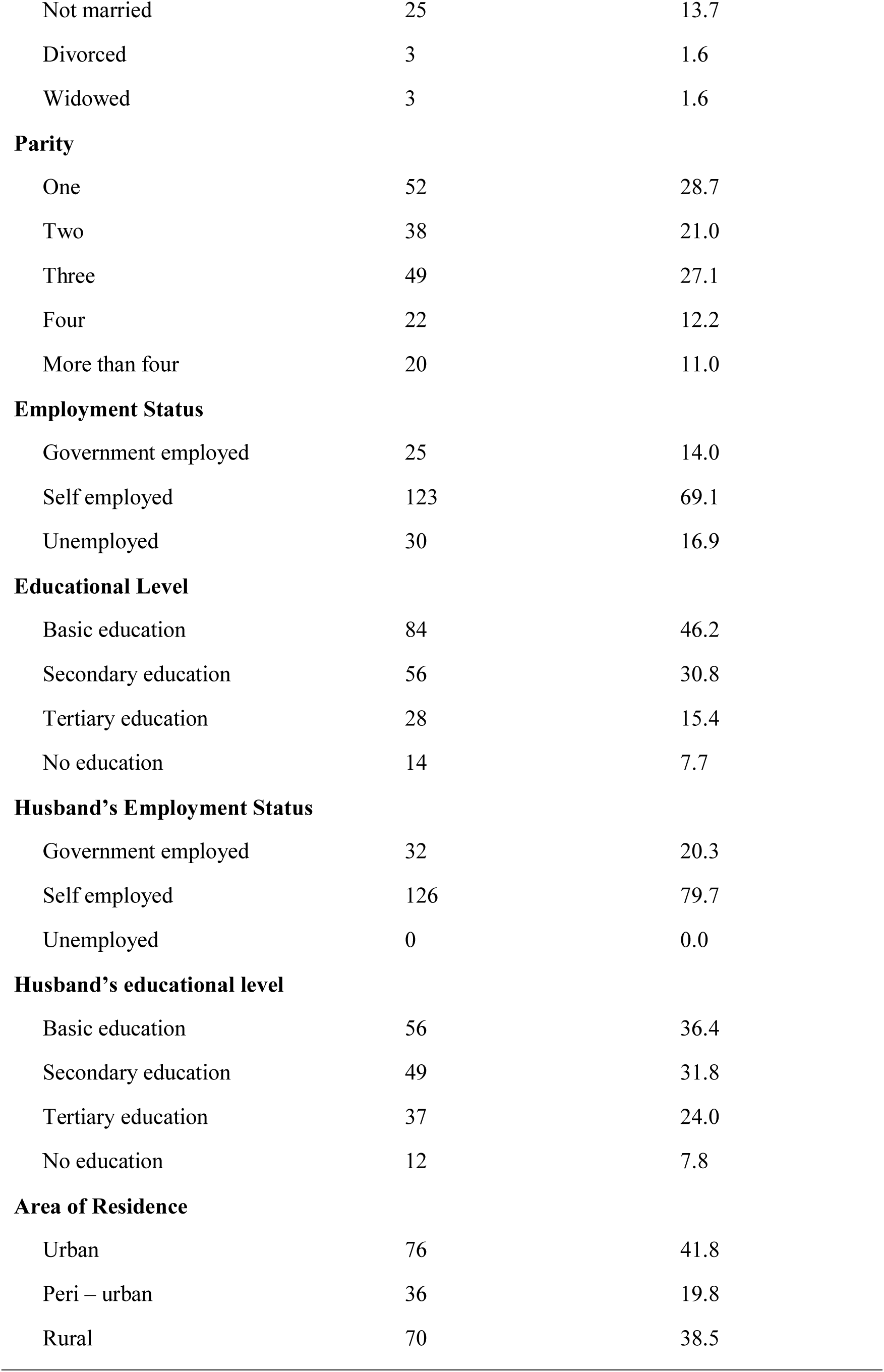
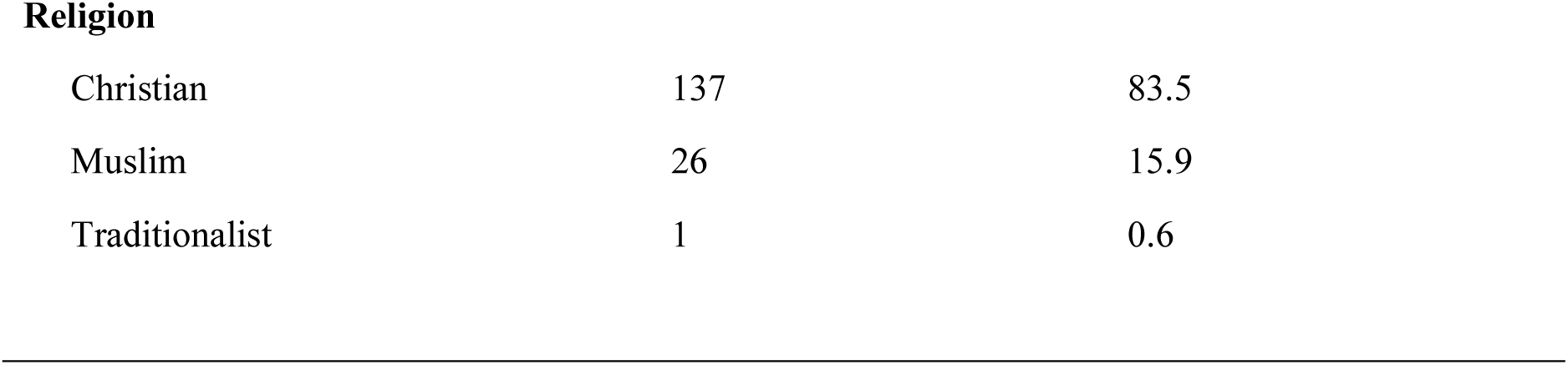
Socio-demographic characteristics of respondents

##### 4.1.2 Antenatal Care Utilization

Antenatal care utilization in this study was measured against three main parameters which included; Antenatal care attendance at least once, number of ANC attendance per WHO recommendation and gestational age at first ANC visit as per WHO recommendation.

###### 4.1.2.1 Antenatal care attendance among postpartum mothers in Sunyani municipality

Data on antenatal care attendance revealed that, a high proportion of women in the Sunyani municipality attended ANC during their last pregnancy birth. Relating the frequency difference, about 87% of the respondents attended ANC during their last pregnancy with only 13% who did not attend ANC during their last pregnancy preceding the study preceding the study.

###### 4.1.2.2 Respondents number of ANC attendance per WHO recommendation

As shown in Fig. 4, majority of the study respondents attained WHO’s recommendation for minimum ANC visit. Among mothers who attended ANC during their last pregnancy, about 95.6% had four or more contacts as recommended by WHO with only a few of them (4.4%) who had less than four contacts.

**Figure 1.0.**
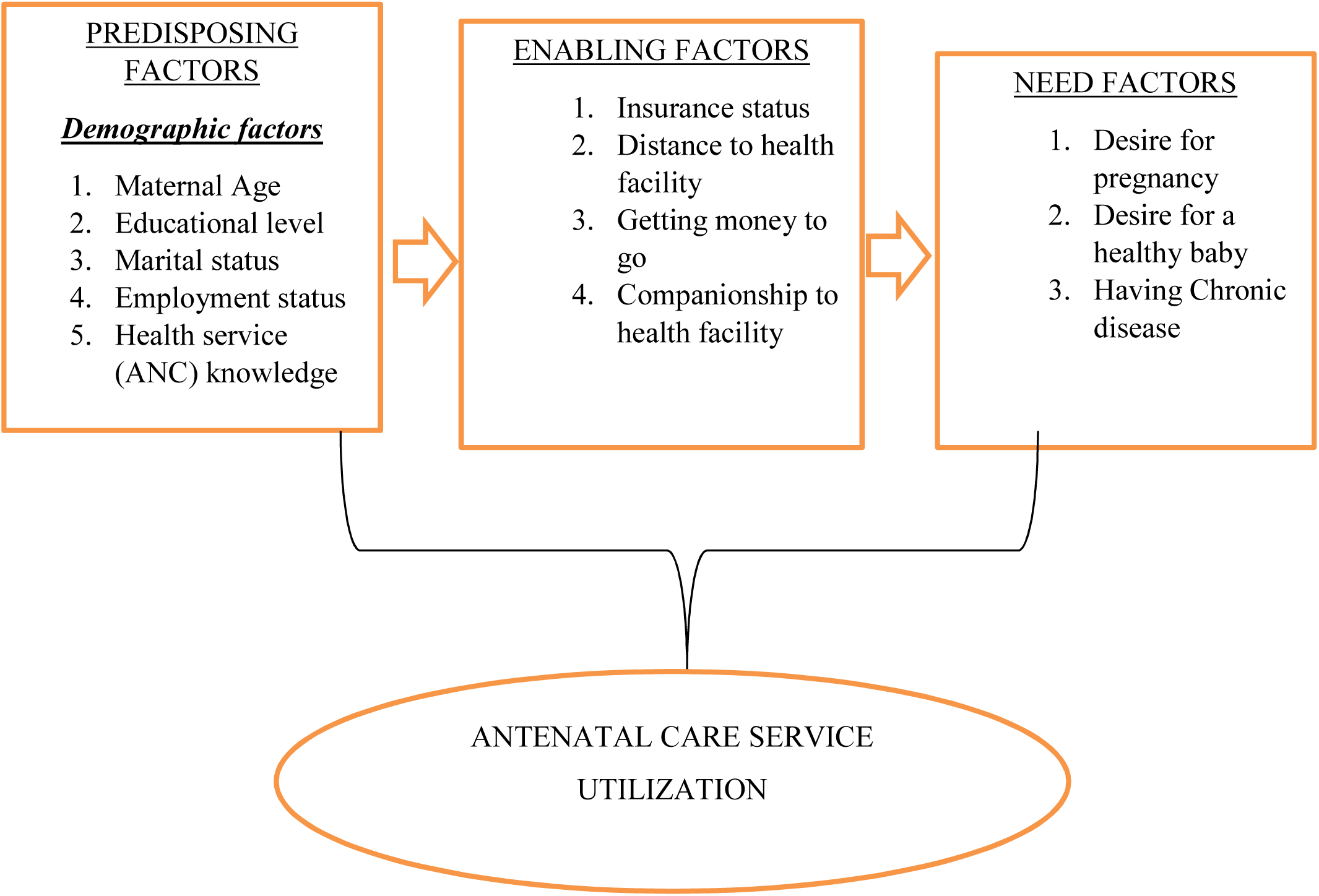
Conceptual framework of factors associated with ANC utilization Source: Andersen and Newman (1973)

**Figure 2.0:**
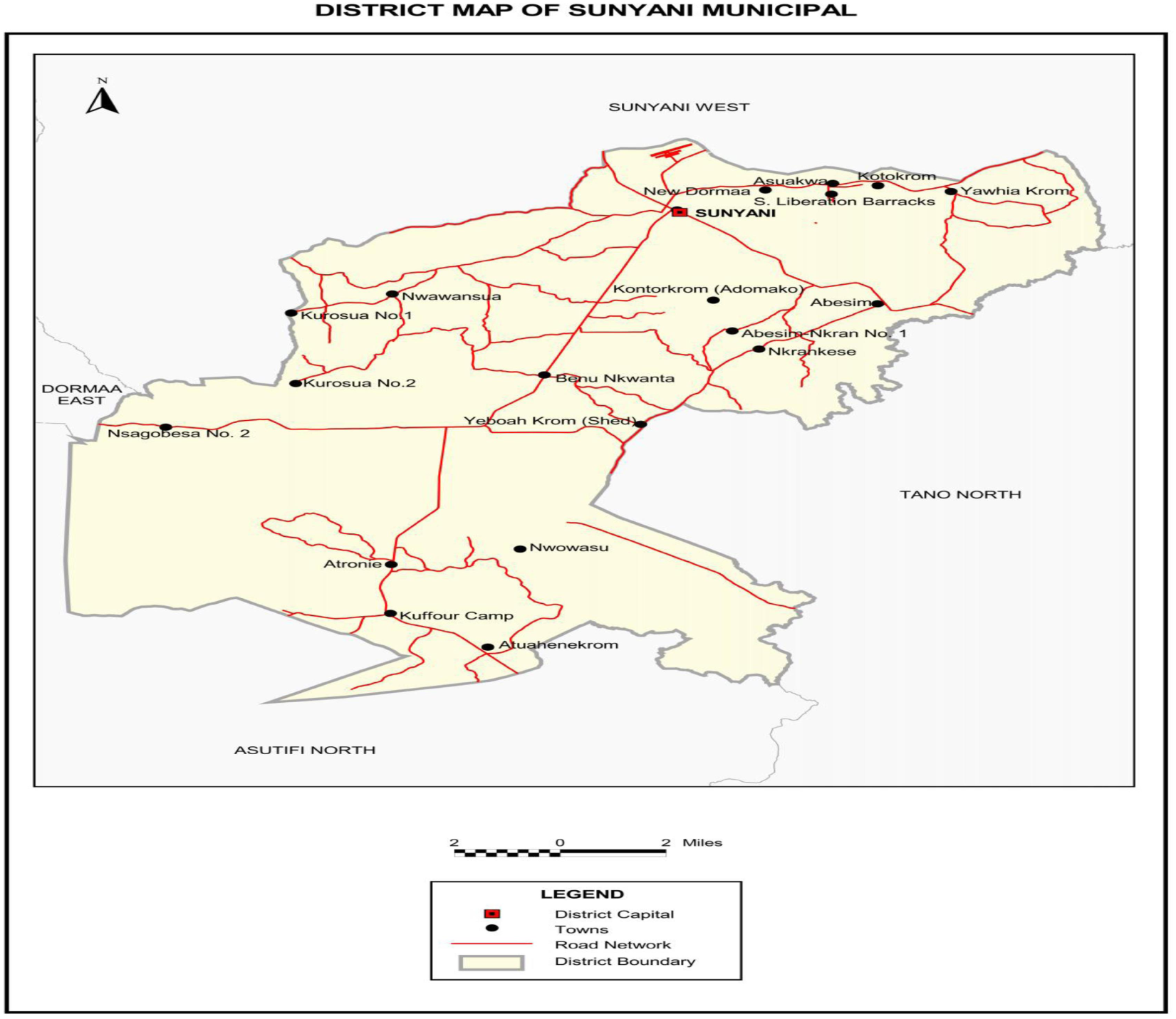
District map of Sunyani municipal. Source: Ghana statistical service, 2014

**Figure 3:**
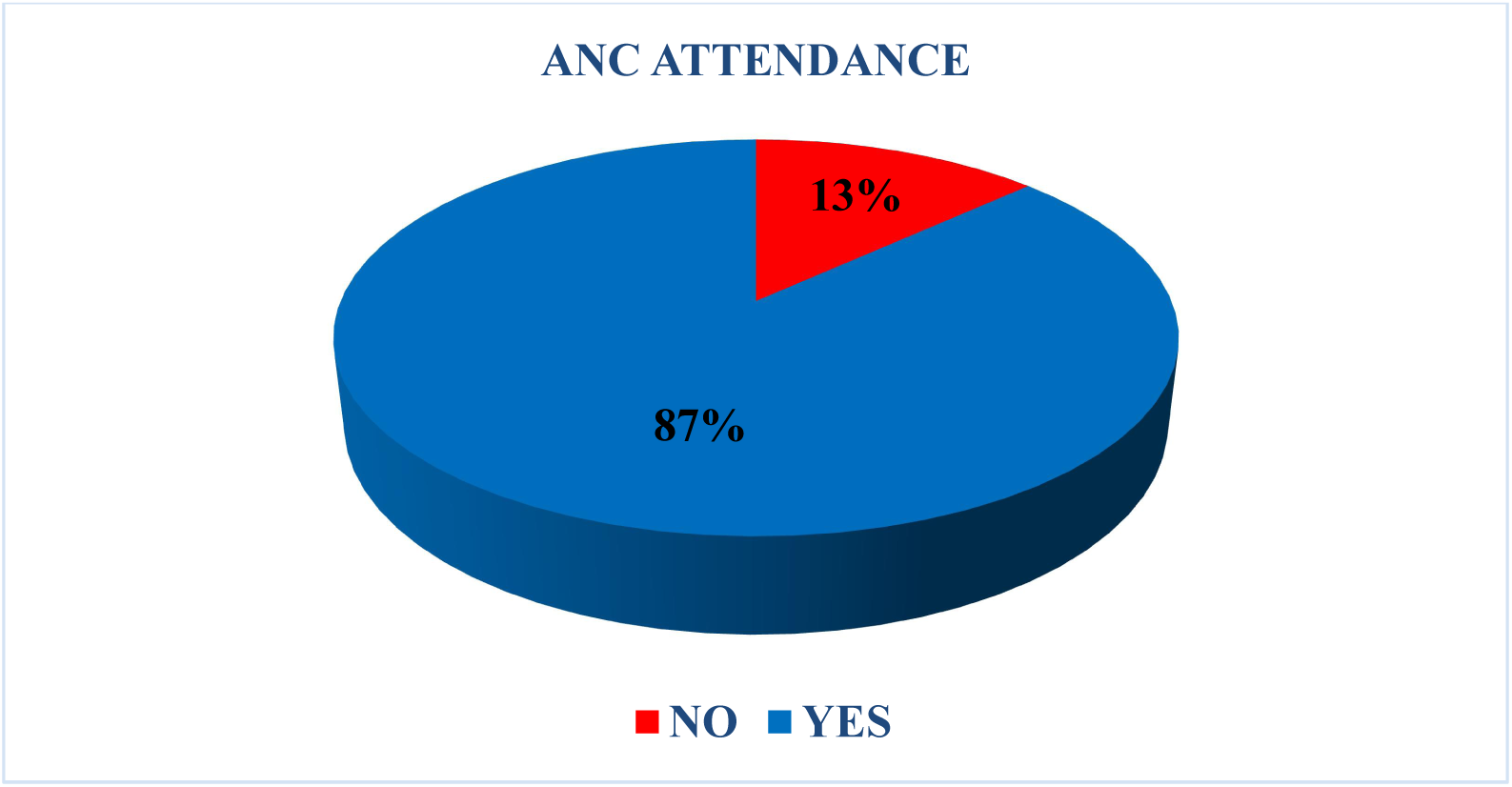
Antenatal care attendance at least once among post-partum mothers in the Sunyani municipality

**Figure 4:**
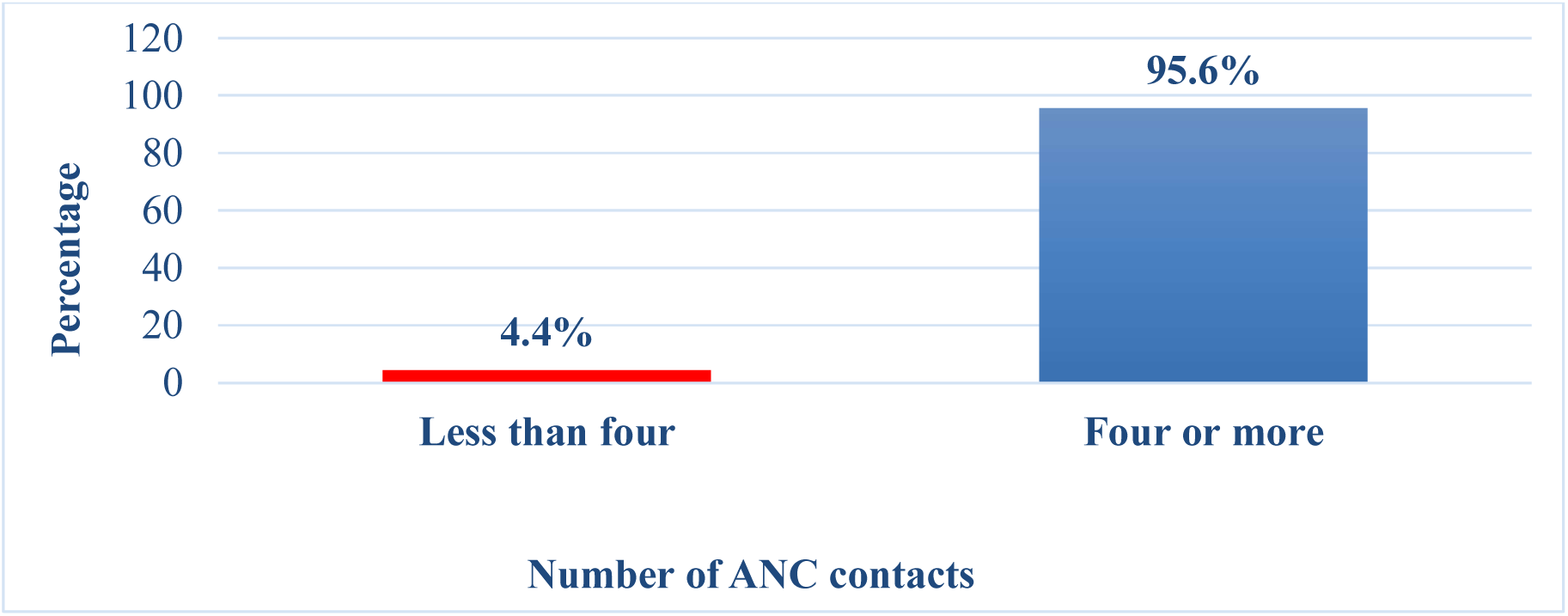
Percentage of respondents who visited ANC per the WHO recommendation

###### 4.1.2.3 Gestational age at which respondents initiated their first antenatal care

Analysis of data on gestational age at which respondents made their first ANC contact measured against WHO recommendation of having it within the first trimester shows that, most of the respondents met this recommendation. About 77.1% of mothers who visited ANC during their last pregnancy had their first contact within their first trimester (within 3moths) as recommended by WHO with few of them (22.9%) having their first ANC contact after their first trimester as clearly depicted in figure 5.

**Figure 5:**
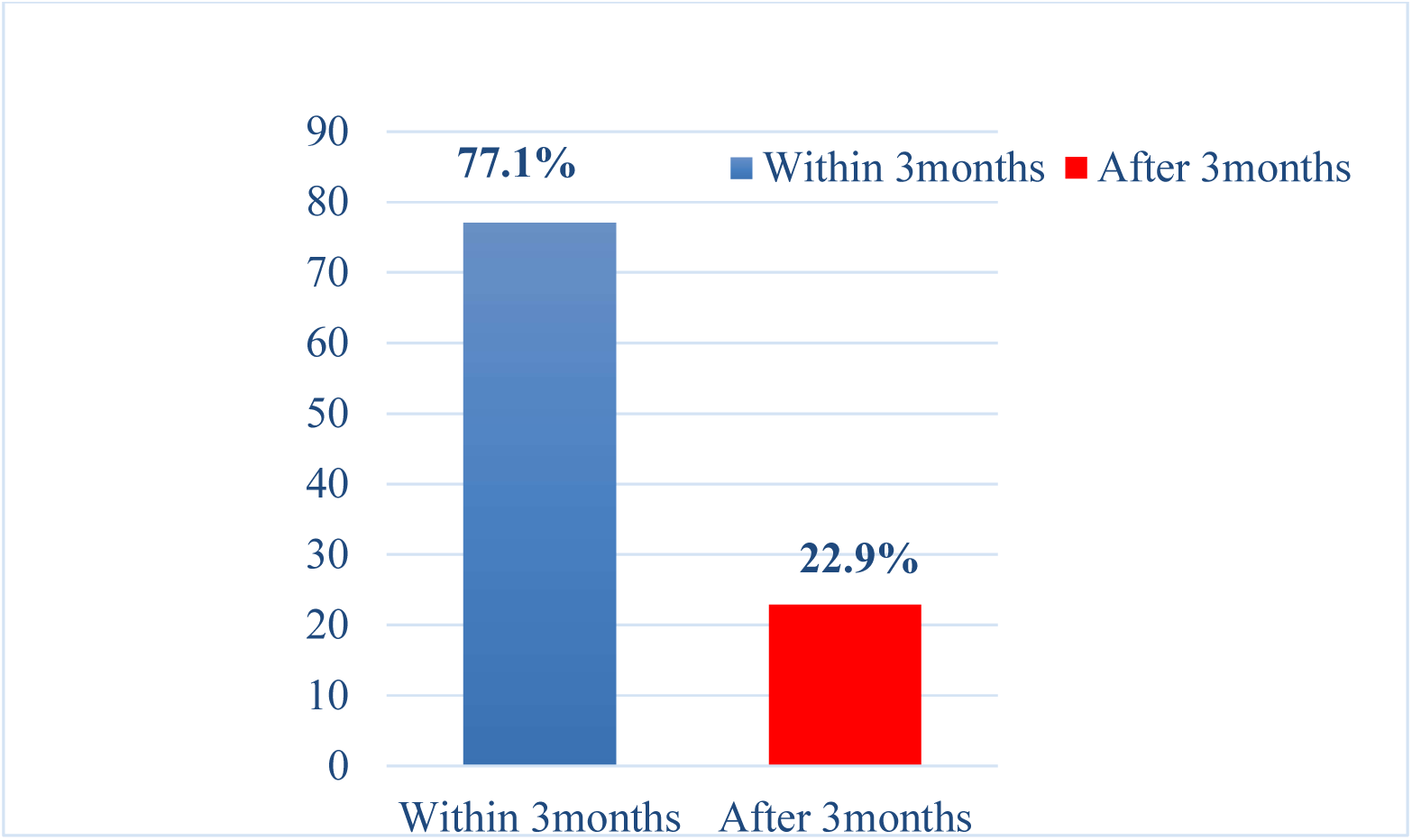
Respondents’ gestational age at first ANC

###### 4.1.2.4 Husbands involvement in ANC attendance in Sunyani municipality

Husbands involvement in attending ANC in Sunyani municipality was observed to be poor. Only a percentage of 16.5 respondents reported that they went to the ANC unit for service with their husbands whiles the majority (79.1%) of the women went alone (figure 6). However, despite husbands not attending ANC with their wives, about 33.5% of them influenced their wives to attend ANC with about 50.6% of the mothers reporting that their attendance to the ANC was not influenced by anybody but they decided to go them self.

**Figure 6:**
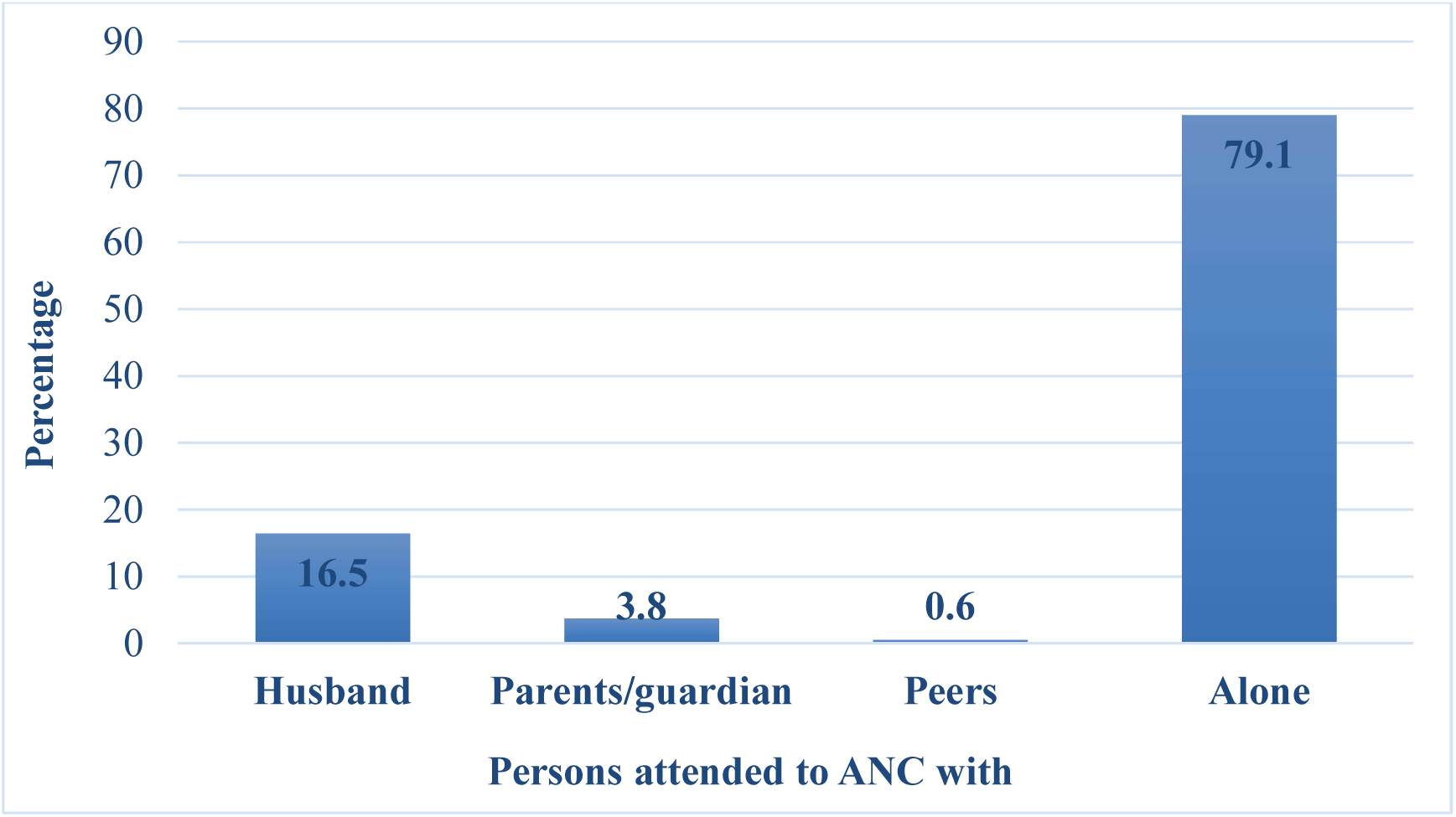
Persons whom respondents went to the ANC with

**Figure 7:**
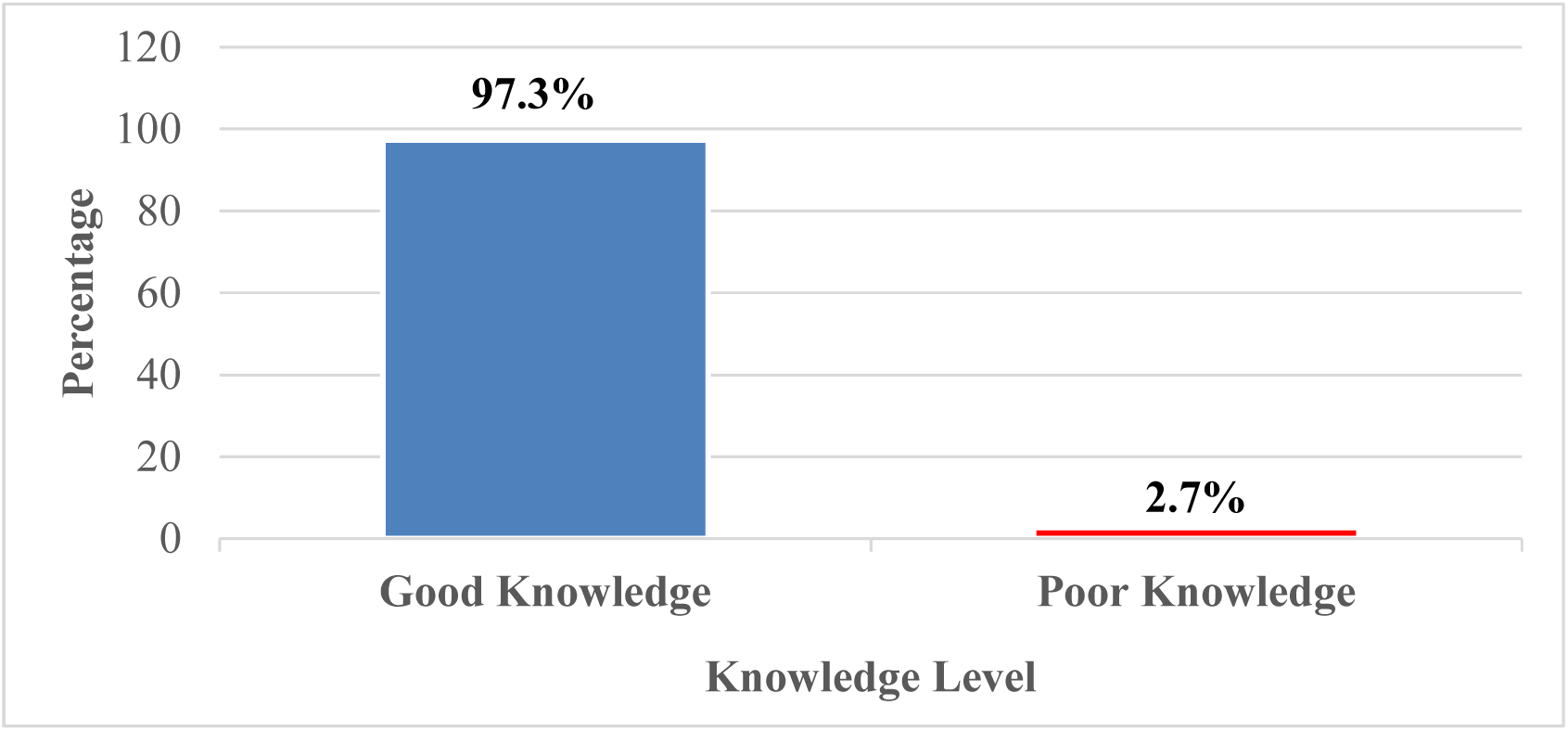
Respondents’ knowledge score about antenatal care services

##### 4.1.3 Bivariate analysis of socio-demographic characteristics associated with antenatal care (ANC) attendance

Analysis for association between socio-demographic features and ANC attendance as presented in table 2 shows that, there was a significant association between age of participants and their ANC utilization, *X^2^* = 19.146, *p* < 0.001. Comparing the percentages, it was found that ANC attendance was highest (94.7%) among respondents’ who are 30 years and above with teenagers (less than 20 years) recording relatively low attendance of about 57.1%. A significant association was also found between respondents’ marital status and ANC attendance, *X^2^* = 26.381^a^, *p* ˂ 0.001. It was however revealed that, majority (93.4%) of the respondents who are married attended ANC during their last pregnancy preceding the study. Almost all the respondents with varied parity (one, two, three, four and ≥ four) had an increased percentage (78.8, 94.7, 91.8, 81.8 and 90.0 respectively) of ANC attendance.

**Table 2:**
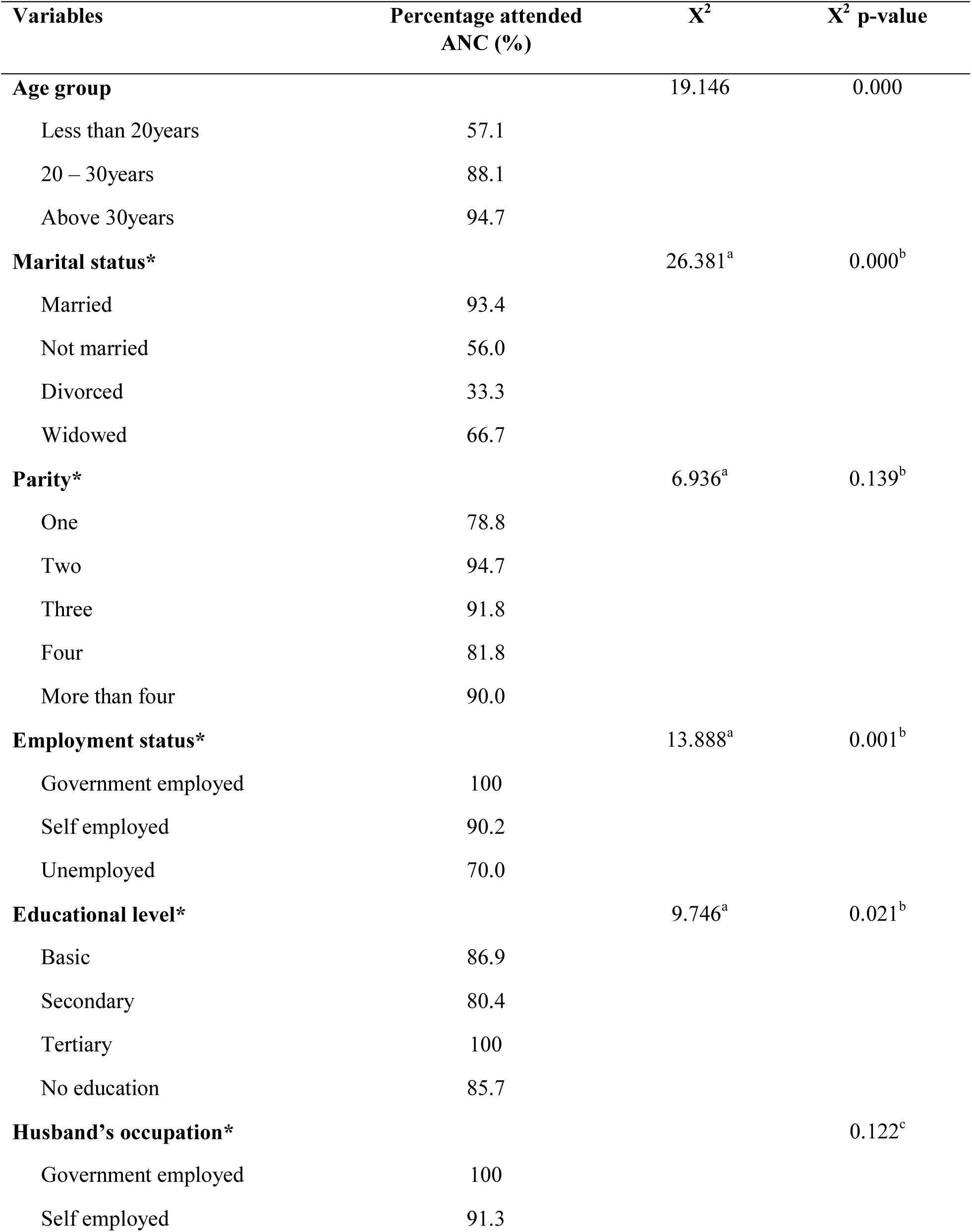
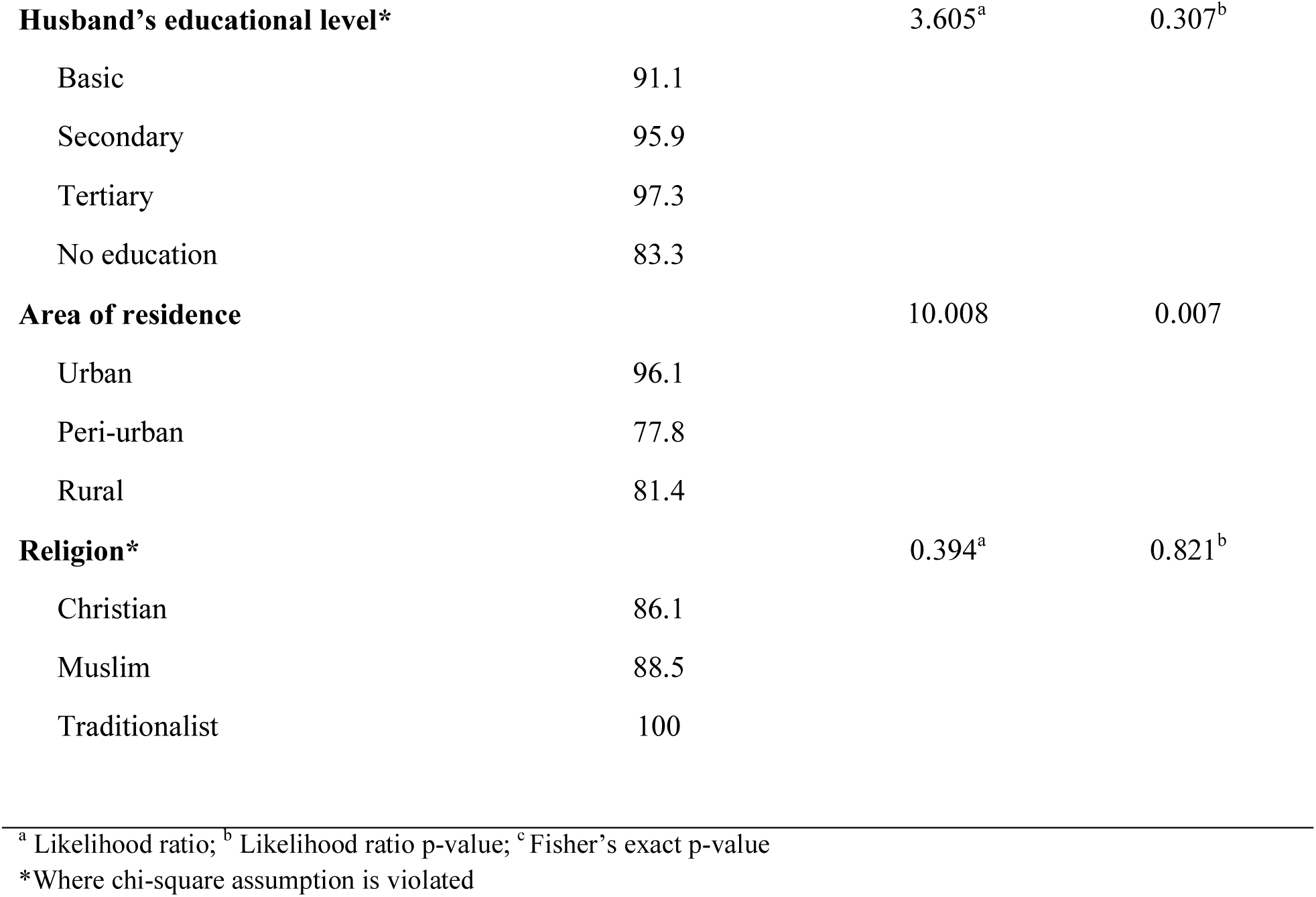
Socio-demographic characteristics of respondents associated with antenatal care attendance

##### 4.1.4 Multivariate analysis socio-demographic characteristics associated with antenatal care (ANC) attendance

In the multivariate logistic regression analysis depicted in table 3, controlling for all possible confounders revealed that, marital status and ANC knowledge were statistically significant with ANC attendance. Respondents who were not married were 78.8% less likely to attend ANC as compared to those married (AOR = 0.212; CI: 0.054 - 0 .834). In addition, respondents who had poor knowledge about ANC were 95.8% less likely to attend ANC using good knowledge as the reference category (AOR = 0.042; CI: 0.002 - 1.113). Employment status and educational level were found not to be statistically associated with ANC attendance during pregnancy. However, age group and area of resident shown significance in the bivariate analysis where mothers who were in the age group of 20 – 30years had 5.56times more odds of attending ANC than those less than 20years or being a teenager and also those residing in rural areas were 82% less likely to attend ANC than those in urban areas but after controlling for confounders, these two variables (age group and area of residence) lost their statistical significance in the multivariate analysis.

**Table 3:** Logistic regression analysis for factors associated with ANC attendance among postpartum mothers in Sunyani municipality.

| **Variable Categories** | **COR (95% CI)**<br>**[P-value]** | **AOR (95% CI)**<br>**[P-value]** |
| --- | --- | --- |
| **Age group** |  |  |
| Less than 20years | Reference | Reference |
| 20 – 30years | 5.563(1.939 - 15.954)<br>[0.001] | 1.416 (0.231 - 8.688)<br>[0.707] |
| Above 30years | 13.50(3.171 - 57.474)<br>[0.00] | 1.692 (0.162 - 17.662)<br>[0.660] |
| **Marital status** |  |  |
| Married | Reference | Reference |
| Not married | 0.090 (0.033 - 0.250)<br>[0.000] | 0.212 (0.054 - 0.834)<br>[0.026] |
| Divorced | 0.035 (0.003 - 0.425)<br>[0.008] | 0.00 (0.00)<br>[0.998] |
| Widowed | 0.142 (.012 - 1.702)<br>[0.123] | 0.118 (0.008 - 1.681)<br>[0.115] |
| **Employment status** |  |  |
| Government employed | Reference | Reference |
| Self employed | 0.000 (0.00)<br>[0.998] | 0.00 (0.00)<br>[0.999] |
| Unemployed | 0.000 (0.00)<br>[0.998] | 0.00 (0.00)<br>[0.999] |
| **Educational level** |  |  |
| Basic education | Reference | Reference |
| Secondary education | 0.616 (0.247 - 1.538)<br>[0.300] | 0.537 (0.166 - 1.738)<br>[0.3] |
| Tertiary education | 2.29e-7 (0.00)<br>[0.998] | 0.00 (0.00)<br>[0.999] |
| No education | 0.904 (0.178 - 4.594)<br>[0.903] | 1.060 (0.130 - 8.630)<br>[0.956] |
| **Area of residence** |  |  |
| Urban | Reference | Reference |
| Peri-urban | 0.144 (0.036 - 0.581)<br>[0.007] | 0.439 (0.084 - 2.279)<br>[0.327] |
| Rural | 0.180 (0.049 - 0.663)<br>[0.010] | 0.244 (0.052 - 1.140)<br>[0.073] |
| **Knowledge score** |  |  |
| Good knowledge | Reference | Reference |
| Poor knowledge | 0.090 (0.014 - 0.569)<br>[0.010] | 0.042 (0.002 - 1.113)<br>[0.023] |

COR – crude odds ratio AOR – adjusted odds ratio CI – confident interval

##### 4.1.5 Respondent’s Antenatal Care Knowledge

Knowledge about ANC service among postpartum mothers was one of the objectives this paper sort to determine in the Sunyani municipality. In table 4 it is indicated that most of the study respondents are having good knowledge about most of the knowledge items that was used for the knowledge assessment. In contrast, about 122 (67.4%) of the study respondents have poor knowledge concerning the minimum number of ANC contacts a pregnant woman should have as per WHO recommendation of four. Respondents included in this study are however seen to have good knowledge (about 97%) about antenatal care service as shown in figure 5.

**Table 4:** Respondents’ knowledge about antenatal care service

| **KNOWLEDGE ITEMS** | **Frequency** | **%** |
| --- | --- | --- |
| ANC should be accessed within the first trimester |  |  |
| YES | 132 | 72.5 |
| NO | 50 | 27.5 |
| Early ANC booking enhances wellbeing |  |  |
| YES | 177 | 97.3 |
| NO | 5 | 2.7 |
| Antenatal care provides the opportunity to detect and manage pregnancy complications |  |  |
| YES | 178 | 97.8 |
| NO | 4 | 2.2 |
| Antenatal care empowers one to be able to identify danger signs and of pregnancy and act promptly |  |  |
| YES | 174 | 95.6 |
| NO | 8 | 4.4 |
| Antenatal care provides an opportunity to learn about STIs, HIV and PMTCT |  |  |
| YES | 175 | 96.2 |
| NO | 7 | 3.8 |
| The minimum ANC contacts a pregnant woman should have is four |  |  |
| YES | 59 | 32.6 |
| NO | 122 | 67.4 |
| Tetanus toxoid immunization and IPTp are among services rendered at the ANC |  |  |
| YES | 172 | 95.6 |
| NO | 8 | 4.4 |

##### 4.1.6 Association Between ANC Knowledge and ANC Service Utilization

###### 4.1.6.1 Bivariate analysis of knowledge and ANC utilization

In table 5 results from cross tabulating knowledge against ANC service utilization shows that, there is a significant association between ANC knowledge and its service attendance, *p* ˂ 0.05. No significant association was found between ANC knowledge and gestational age for first ANC visit as well as between ANC knowledge and number of visits, *p* ˃ 0.05. About 98.7% of respondents who has good knowledge about ANC actually attended ANC during their last pregnancy preceding the study where about 87.5% of those with good knowledge not attending ANC. As opposed to those with poor knowledge score, only 1.3% attended ANC during their last pregnancy preceding the study.

**Table 5:**
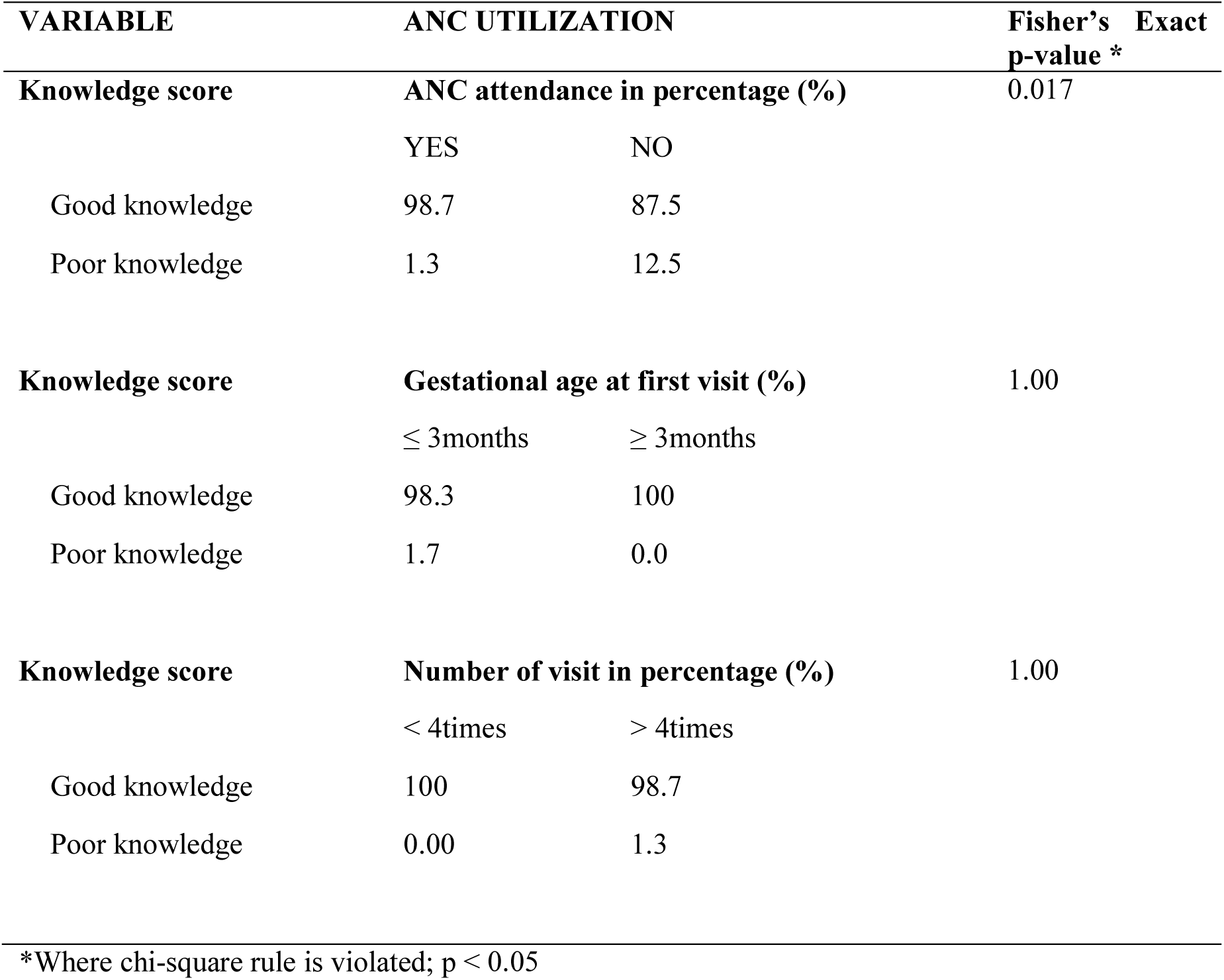
Respondent’s knowledge score and association with ANC utilization

#### 4.2 Discussions

##### 4.2.1 Introduction

This study was aimed at determining factors associated with antenatal care service utilization in Sunyani municipality. This chapter discusses the result of the study in relation to the objectives and key variables of the research.

##### 4.2.2 Antenatal care service utilization

Antenatal care service utilization in this study was measured against three parameters to estimate how well postpartum mothers in the Sunyani municipality use ANC before birth. These parameters included; whether or not respondent attended ANC during her last pregnancy, the number of attendance per WHO recommendations and gestational age at first ANC visit.

Findings from this study shows that majority (87%) of mothers who have recently delivered during the period of this study attended ANC at least once before delivery. Most of these mothers (95.6%) had the WHO recommended number of visits (4 or more) and about 77% of those who attended ANC initiated their attendance within the first trimester. This could be due to how important respondents sees ANC services and the value they place on the service as majority of the mothers (94.3%) reported that they decided to attend ANC because they wanted to prevent pregnancy complications. Most (86.0%) wanted to have a healthy baby with a few (1.3%) been forced to go. The relatively high rate of ANC attendance observed in the Sunyani municipality is found to be in agreement with some recent literatures. For example, a study by Abubakari and Abiiro (2018) found an increased ANC visit (81%) with a coverage of 99% in Northern Ghana. Also between 2011 and 2017, almost all pregnant women in Lower Middle Income Countries (LMICs) paid a visit to the ANC at least once (Tikmani et al., 2019). In addition, Adu et al. (2018) also observed that, many Ghanaian women (89.2%) attended ANC during pregnancy four or more with the few (10.8%) having less than four visits. Yamba (2018) also adds it on that, about 98% of Sierra Leone women attended ANC at least once with 89% having four (4) or more visits.

However, these findings are found not to be in agreement with some other literatures. Taking a study by Adewuyi et al. (2018) for example, the prevalence of ANC underutilization in Nigeria was found to be 46.5%. In addition, findings from Amoah et al. (2016) shows that, only 31.7% of their respondents had their first ANC visit within the first three months. Moreover, only 25% of pregnant women in Rwanda begins their ANC visit within the timeframe recommended by WHO (Mkandawire et al., 2019).

In addition, this study found male partners’ involvement in ANC to be poor as only 26 (16.5%) of mothers reported that they went to the ANC with their husband whiles majority 125 (79.1%) of the mothers reported they went alone. In support of this finding, Ganle and Dery (2015) explored in their study that, men involvement in maternal health issues is only ignited when there is a complication during pregnancy or child birth if not, they never involve them self in maternal health issues although they know its importance. This could be as a result of gender norms and gender roles which persist in some societies creating the perception that, women are solely responsible to ensure healthy pregnancy whiles men are family providers or breadwinners. However, a study by Tweheyo, Konde-Lule, Tumwesigye, and Sekandi (2010) challenges this finding as they observed that, 65% of men in Northern Uganda participated at least once in ANC services with an increased willingness (93.7%) to attend subsequent ones.

##### 4.2.3 Predisposing factors of demographic characteristics associated with ANC attendance

Many literatures have identified a number of factors which predisposes pregnant mothers’ intention to attend or not to attend ANC. Among such factors include demographic characteristics of the pregnant mother. One of the objectives of this study was to determine which demographic factors are associated with ANC attendance among postpartum mothers in Sunyani municipality.

Results from this study shows that, marital status and knowledge about antenatal care services are significant predictors of ANC attendance. For marital status, mothers who were not married were about 78.8% less likely to attend ANC as compared to those who were married (AOR: 0.212; CI: 0.054 - 0 .834). These findings corroborate the results of a study by Sakeah et al. (2017). They found out that, cohabiting women and unmarried women were 43% and 61% respectively less likely to attend ANC at least four times relative to married women. Moreover, findings from Ziblim, Yidana and Mohammed (2018) affirms that there is a significant association between marital status and ANC utilization. Furthermore, it has been found that women who are single, divorced, widowed or separated stands a higher chance of poor ANC utilization compared to married women (Rurangirwa et al., 2017).

This could be due to certain social norms defined in some societies and cultures including Ghana where unmarried women are expected to remain chaste until they are married (Malacane & Beckmeyer, 2016). Therefore, unmarried women who becomes pregnant are more likely to avoid ANC services so as not to expose their pregnancy for fear of public ridicule. In addition, women who do not have partners could experience financial difficulty that might prevent them from attending ANC regularly.

In variance to these findings, Nuamah et al. (2019) found no significant association between marital status and ANC utilization with those cohabiting having a higher odds of attending ANC compared to those who are married. Also findings from Kparu (2016)’s study in Ghana suggest no significant association between marital status and ANC attendance.

Employment status and maternal education were variables found not to be statistically significant with ANC attendance in this study. Some literatures however approve on this observation. For example, according to Adu (2018), there is no association between maternal education and ANC attendance. In contrast, some studies also reject this observation. For instance, a study by Adewuyi et al. (2018) found maternal educational level and wealth index as factors which predicted ANC attendance in Nigeria. Again, maternal health service utilization in general is found to be associated with maternal education and wealth quantile according to a study in Eritrea (Habtom, 2017).

##### 4.2.4 Antenatal care service knowledge and its prediction for ANC attendance

Findings from this study revealed that, all most all (97%) of the study respondents are having good knowledge about antenatal care services. Although 122 (67.4%) of the respondents did not know the exact minimum number of ANC contacts a pregnant woman should have as per WHO recommendation of four contacts throughout her pregnancy period, this knowledge gab did not have any negative effect on the number of visits respondents had during their last pregnancy as some of the respondents reported that four contacts are not enough for healthy pregnancy. It was again noted in this study that, respondents had good knowledge as to when a pregnant woman should initiate ANC. This resulted in majority (77%) of them initiating their ANC attendance within the recommended gestational period (within first three months). But the findings from Respress et al. (2017) challenges this observation as they found that, some percentage of women (24.6%) in Jamaica are not aware of when a pregnant woman should initiate her first ANC.

In addition to this present study, majority of the respondents had good knowledge about some of the important provisions of ANC. This could however be a reason for the high ANC coverage and or attendance that was observed as they were influenced to attend ANC by been aware of its tremendous benefits. In a study done in Manipur, similar findings were observed where majority (97.9%) of postpartum mothers were aware that pregnant women have to attend ANC before birth (Laishram, ThounaojamUd, Mukhia, & Sanayaima, 2013). However, Kaur and colleague (2018)’s study findings debate against the fact that most women are not aware about the minimum number of ANC a pregnant woman should have. In their present study, they observed that 89% of their respondents were able to tell the correct number of minimum ANC visits a pregnant woman should have in the course of her pregnancy till birth.

As part of this study’s objectives, the association between knowledge and ANC attendance was aimed to be known. It was however found that, the level of mother’s knowledge on ANC services was found to be significantly associated with ANC service attendance. Mothers with poor knowledge were about 95.8% less likely to attend ANC compared to mothers with high knowledge about ANC services. This finding is therefore been buttressed by a lot of current literatures. Taking a study by Respress and colleagues (2017) for example, mothers with good knowledge attended ANC not less than four times as against those with poor knowledge, generating an association between these two variables (knowledge and utilization). Also according to Ogboghodo, Adam, Omuemu and Okojie (2019), good knowledge on maternal health issues can be deemed as a clairvoyant for skill birth attendance (SBA). Moreover, Mwilike et al. (2018) also have established an association between knowledge on danger signs of pregnancy and the willingness to seek healthcare assistance in Tanzania. Yet, Ogunba and Abiodun (2017) is found to be in opposition to this finding by way of finding no significant relationship between knowledge and the attendance to antenatal clinic in a study they conducted.

## CHAPTER FIVE

### 5.0. CONCLUSIONS AND RECOMMENDATIONS

#### 5.1 Conclusion

It has been revealed by this study that, majority of postpartum mothers in the Sunyani municipality utilized antenatal care during their last pregnancy before birth. The utilization included increased number of ANC attendance (more than 4times) and early initiation (within the first trimester). These postpartum mothers were again observed to have an increased knowledge about antenatal care services with the exception of few whom were not aware about the minimum number of ANC contacts a pregnant woman should have before birth.

In addition, marital status and ANC knowledge were found to be factors which predisposes women either to use or not to use ANC services.

Finally, husbands were observed to be poorly involved in the attendance of antenatal care in the Sunyani municipality of Ghana.

#### 5.2 Recommendations

Based on these findings, it is recommended that;

1. Health education programs should be initiated that target men in order to create awareness about the importance of ANC services and why they should involve themselves as husbands and strengthening/implementing incentives for husbands who involve themselves in ANC.
2. Health promoters and educators should include the minimum and maximum ANC contact pregnant women are supposed to have according to the WHO recommendations in their education and promotion sessions.
3. Another study can be done focusing only on adolescence to explore the factors associated with their ANC and other maternal health service utilization.

## Data Availability

All data presented or referred to in the manuscript is available

## LIST OF ACRONYMS

ANC: Antenatal Care
CHAG: Christian Health Association of Ghana
CHPS: Community-based Health Planning Services
CWC: Child Welfare Clinic
DHMT: District Health Management Team
EPI: Expanded Programs on Immunization
GHS: Ghana Health Service
GSS: Ghana Statistical Services
HBM: Health Belief Model
HMT: Hospital Management Team
IPTp: Intermittent Preventive Therapy for pregnant women
MDG: Millennium Development Goal
MMR: Maternal Mortality Ratio
MoH: Ministry of Health
SDG: Sustainable Development Goal
UI: Uncertainty Interval
UNFPA: United Nations Population Fund
UNICEF: United Nations International Children’s Emergency Fund
UNPD: United Nations Population Division
WHO: World Health Organization

## APPENDIX 1: COPY OF ETHICAL CLEARANCE CERTIFICATE

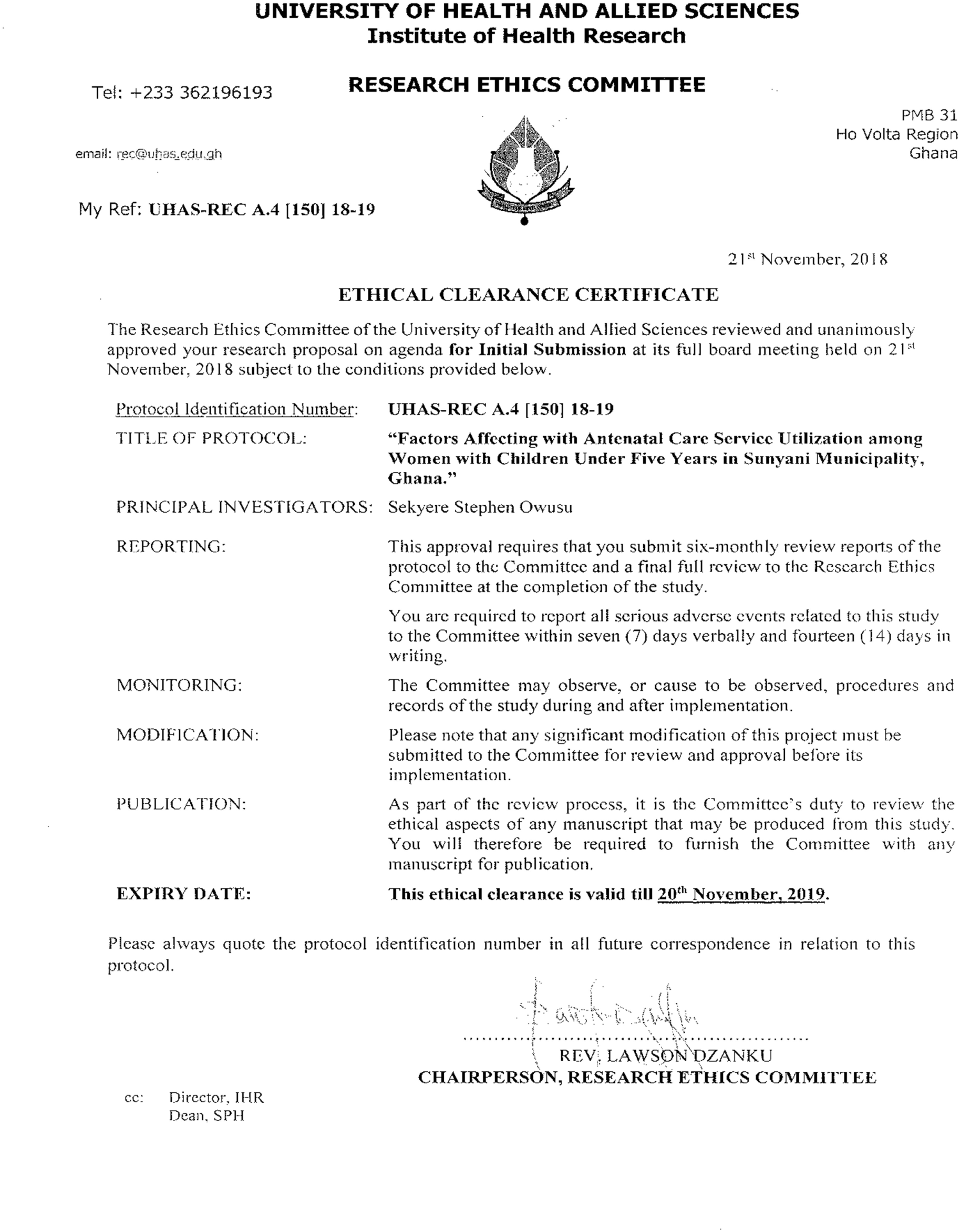

